# High secondary attack rate and persistence of SARS-CoV-2 antibodies in household transmission study participants, Finland 2020

**DOI:** 10.1101/2021.07.25.21260925

**Authors:** Timothée Dub, Hanna Nohynek, Lotta Hagberg, Oona Liedes, Anu Haveri, Camilla Virta, Anna Solastie, Saimi Vara, Nina Ekström, Pamela Österlund, Katja Lind, Hanna Valtonen, Heidi Hemmilä, Niina Ikonen, Timo Lukkarinen, Arto A. Palmu, Merit Melin

## Abstract

**Background:** Household transmission studies offer the opportunity to assess both secondary attack rate and persistence of SARS-CoV-2 antibodies over time.

**Methods:** We invited confirmed COVID-19 cases and their household members to attend up to four household visits with collection of nasopharyngeal and serum samples over 28 days after index case onset. We calculated secondary attack rates (SAR) based on the presence of SARS-CoV-2 nucleoprotein IgG antibodies (IgG Ab) and/or neutralizing antibodies (NAb) overall and per households. Three and six months later, we assessed the persistence of SARS-CoV-2 antibodies.

**Findings:** We recruited 39 index cases and 90 household members. Among 87 household members evaluated, SAR was 48% (n=42), including 37 symptomatic secondary cases. In total, 80/129 (62%) participants developed both IgG Ab and NAb, while three participants only developed IgG Ab. Among participants who had both IgG Ab and NAb during the initial follow-up, 68/69 (99%) and 63/70 (90%) had IgG Ab and NAb at 3 months, while at 6 months, 59/75 (79%) and 63/75 (84%) had IgG Ab and NAb, respectively. Participants who required hospital care had initially 5-fold IgG Ab concentrations compared to cases with mild symptoms and 8-fold compared to asymptomatic cases.

**Interpretation:** Following detection of a COVID-19 case in a household, other members had a high risk of becoming infected. Follow-up of participants showed strong persistence of antibodies in most cases.

**Funding:** This study was supported by THL coordinated funding for COVID-19 research (Finnish Government’s supplementary budget) and by the Academy of Finland (Decision number 336431).

**Research in context:** *Evidence before this study:* Household transmission studies are pivotal to the characterization of transmission dynamics of emerging infectious diseases in a closed setting with homogenous exposure, including proportion of asymptomatic cases using serologic assessment of infection. Additionally, data on long-term persistence of immune response, including neutralizing antibodies following COVID-19 remains scarce. Our search on PubMed for articles published between January 1^st^ 2020, and June 1^st^, 2021 using the search terms “household” AND “transmission” AND (“COVID-19” OR “SARS-CoV-2”) retrieved 381 results including 35 relevant articles: 21 original household transmission studies, 5 reviews and 9 statistical transmission, modelling or register linkage studies. Depending on the diagnosis method and the duration of follow-up, secondary attack rates (SAR) ranged from 4.6% when household contacts were followed for 14 days and tested only in case of symptoms to close to 90%. None of the household transmission studies involved long-term convalescent follow-up.

*Added value of this study:* This extensive (one month) active follow-up, using RT-PCR diagnosis and serological testing for SARS-CoV-2 nucleoprotein IgG antibodies (IgG Ab) and neutralizing antibodies (NAb) showed that household transmission was high, with a 48% (42/87) SAR overall and 50% [IQR: 0-100%] at the level of the household. All but one out of 64 RT-PCR confirmed participants had developed both IgG Ab and NAb after immediate convalescence. Six months after inclusion, majority of previously seropositive (IgG and/or NAb) participants still had IgG Ab (59/75) or NAb (63/75) showing long-term persistence of humoral immunity to SARS-CoV-2.

*Implications of all the available evidence:* The risk of transmission of SARS-CoV-2 infections within households is considerable. Isolation of the primary case, especially from household contacts with a high risk of severe disease, e.g. due to age or comorbidities, should be considered even though viral shedding might occur before confirmed diagnosis in household contacts. Long-term persistence of antibodies following infection, even in asymptomatic and mild cases, suggests enduring natural immunity and possibly protection from severe COVID-19.

## Introduction

In December 2019, SARS-CoV-2, a novel coronavirus causing COVID-19 was detected in the city of Wuhan (China).^1^ Its emergence rapidly led to a pandemic and more than 190 million confirmed cases, including four million deaths have been reported.^2,3^ As with any newly detected respiratory pathogen, the emergence of SARS-CoV-2 was associated with a lack of knowledge of several key parameters in infectious disease epidemiology, including transmission dynamics, the expected severity of the infection, the development of antibodies and duration of immunity.

In Finland, the first locally acquired case was detected in February 2020 and public health measures to control transmission were implemented on March 12, 2020 including a recommendation for remote working and a ban on events and gatherings with more than 500 participants, before a state of emergency (closure of schools, universities and public places, limit of ten person per gatherings, recommendation to reduce social interactions) was declared on March 16, 2020.^4^ We undertook a SARS-CoV-2 household transmission study in the Helsinki capital area and followed study participants for a duration of 6 months to assess long-term persistence of SARS-CoV-2 specific antibodies.

## Methods

### Study design and participants

#### Household transmission

We conducted a household transmission study starting in March 2020 using the Household transmission investigation protocol for coronavirus disease 2019 (COVID-19) developed by the World Health Organisation.^5^ We invited a convenience sample of cases with a recent PCR-confirmed SARS-CoV-2 infection and their household members residing in the Helsinki capital area to participate in four household visits at day 0, day 7, day 14 and day 28 after onset of the index case. Participants were either recruited after invitation by a text message obtained during the contact enquiry of the index case or COVID-19 cases could directly be enrolled after reading the study information on the institute’s website.^6^ At each of the four visits, we collected blood and nasopharyngeal samples of all index cases and household members to detect infections by PCR and to assess SARS-CoV-2-specific antibodies induced by infection. We also collected data from index cases and contacts on symptoms and their date of onset, as well as a symptom diary collecting history of symptoms and their onset for 28 days following onset of the index case.

#### Follow-up of SARS-CoV-2 immunity at three and six months

We invited cases that had developed antibodies during the initial 28-day follow-up for serum collection at 3 months post onset of the index case and all study participants for serum collection at 6 months post onset of the index case.

#### Sample collection techniques

To detect acute SARS-CoV-2 infection at any point in the follow-up period, we collected a nasopharyngeal specimen from all household members at each visit. The nasopharyngeal specimens were stored in a transport medium immediately after the sampling and transported at ambient temperature to the reference laboratory at THL for testing by RT-PCR. To detect antibodies induced by SARS-CoV-2 infection, we collected a venous blood sample from each household study participant into a 10/8.5 mL BD Vacutainer gel serum tube containing clot activator (reference number: 367955) during the initial follow-up and three months visit. At six months, we used BD Vacutainer 5/3.5 ml gel serum tube with clot activator (reference number: 367957) and a smaller tube 5/2 ml (reference number: 368492) for children. When collection of a venous samples was not possible, we collected a fingertip sample into a BD Microtainer serum microtube with clot activator (reference number: 365964).

#### Microbiological analyses

##### Reverse transcriptase Polymerase Chain Reaction (RT-PCR)

We extracted RNA from specimens using the Qiagen Qiacube^R^ instrument with RNeasy Mini Kit^R^. We synthesized cDNA using random hexamer primers and RevertAid H Minus Reverse Transcriptase and tested specimens in three separate real-time polymerase reaction tests using QuantiTect^™^ Multiplex NoRox PCR Kit. Primers and probes were targeted for the envelope (E), the RNA dependent RNA polymerase (RdRp) and the nucleocapsid (N) genes. The sequences of primers and probes are published in Corman et al.^7^

##### Serologic investigations

###### Fluorescent microsphere immunoassay

We used an in-house fluorescent microsphere immunoassay (FMIA) for measurement of IgG antibodies (IgG Ab) to SARS-CoV-2 nucleoprotein. We have previously described the validation of the assay. Briefly, sera were diluted to 1/100 and 1/1600, in-house reference serum (1/400 to 1/1 638 400) and two control sera per plate were mixed with SARS-CoV-2 nucleoprotein conjugated fluorescent microspheres and IgG antibodies were detected by R-Phycoerythrin (RPE) -conjugated secondary antibody and read the plate on MAGPIX® system using xPONENT software v4.2 (Luminex®Corporation, Austin, TX). The in-house reference sample was assigned an arbitrary IgG antibody concentration of 100. Median fluorescent intensity (MFI) was converted to FMIA unit/ml (U/ml) by interpolation from a 5-parameter logistic (5-PL) reference curve. Using a 1.71 U/ml cut-off resulted in 100% specificity and 100% sensitivity.^8^

###### Microneutralization test

To measure SARS-CoV-2-specific neutralizing antibodies (NAb), we used the cytopathic effect (CPE)-based microneutralization test (MNT) as previously described.^9^ Serum samples were analysed in duplicate, 2-fold serial dilution starting from dilution 1:4. We analysed all samples with two viruses isolated from COVID-19 patients in Finland in the spring 2020; hCoV-19/Finland/1/2020 (GISAID accession ID EPI_ISL_407079): Fin1-20 and hCoV-19/Finland/FIN-25/2020 (EPI_ISL_412971): Fin25-20. A positive control sample from a convalescent COVID-19 patient was included in each analysis to assess inter-assay variation. For Fin1-20 strain, we obtained a MNT titer of 192 for the NIBSC-standard 20/136.^10^ If both MNT titers were <4, or if titers were <4 and 4, a sample was considered as negative. When MNT titers were equal to 4 with both viruses we considered the sample as borderline. Finally, a sample with a MNT titer ≥6 with any of the viruses was considered as positive. Samples with negative titers were given a value of 2 for statistical analysis.

We analysed the serum samples collected during the household transmission study, and during the follow-up at three months ongoingly soon after sample collection. Due to the inter-assay variation in the MNT, NAb titers can be reliably compared at an individual level only when the samples are analysed within the same batch.^8^ Therefore, the convalescent samples collected at the six month follow-up were analysed with the previously collected serum samples for each patient at one month and three months in order to describe trends of NAb titers on an individual level.

### Operational definitions

We defined a household as a group of two people or more living in a domestic residence, and a household contact as a person who resided in the same household as the index COVID-19 case while symptomatic. We excluded collective residence such as boarding schools, hostels or prisons. For inclusion of a household, at least one member had to have a recently confirmed SARS-CoV-2 infection by RT-PCR through a routine healthcare contact. This household member was considered as the index case. We defined a case as a participant who either had RT-PCR confirmation of SARS-CoV-2 infection, or who had either developed IgG Ab or NAb by the end of the initial follow-up.

### Statistical analysis

#### Household transmission

##### Reclassification of index and secondary cases and exclusion of households

In households where the index case was the first one to have experienced symptoms, index cases were considered as primary cases. If a secondary case reported earlier symptom onset than the index case, then the secondary case was reclassified as primary case, and the index case as a secondary case. If onset of symptoms was the same for the index and at least one household secondary case, then this household was excluded from the household transmission analysis but still included in the long-term serological follow-up.

##### Descriptive analysis

We calculated the number of secondary cases overall and per households as well as the effective household reproduction number depending on the age of the primary case. We described healthy household members and secondary cases characteristics: age, sex, pre-existing conditions and symptoms using Chi2 or Fischer exact test when sample size required.

#### Assessment and follow-up of SARS-CoV-2 immunity

We used geometric mean concentrations (GMC) and titers (GMT) and their 95% confidence intervals, for IgG Ab and NAb, respectively, to describe trends in antibody concentrations and compared antibody levels of cases that had required hospital care to cases with mild disease and cases with asymptomatic infection. We also assessed the persistence of antibodies in the initially seropositive cases over time. We performed statistical analysis using STATA 15.1 (StataCorp LP Lakeway, TX, USA).

### Ethics

The Finnish communicable diseases law and the law on the duties of the Finnish Institute for Health and Welfare allowed the implementation of the initial household transmission study and 3 months convalescent sample collection without seeking further institutional ethical review^11,12^. The protocol for the follow-up visits at 6 months was reviewed and approved by the HUS ethical committee and registered under the Development of seroprevalence in Finland during the new coronavirus (SARS-CoV-2) epidemic – serological population study protocol HUS/1137/2020. Informed consent was obtained from all cases and contacts or their parents or legal guardian before any procedure was performed.

## Results

Between March 24^th^ and June 17^th^ 2020, we recruited 39 index cases and 90 household members, i.e. a total of 129 participants from 39 households. Overall, 84 (65%) participants had evidence of SARS-COV-2 infection, confirmed either by PCR-testing (n=64, 50%) or serology (n=83, 64%). Of the seropositive cases, 83 and 80 developed IgG Ab or NAb, respectively. Out of 84 COVID-19 cases, five did not experience any symptoms, 75 had experienced mild symptoms, while four required hospital care (Table 1).

**Table 1:** Description of household study participants.

|  | Primary cases<br>(n=37)<br>N (%) | Household contacts<br>(n=87)<br>N (%) | Excluded participants<br>(n=5)<br>N (%) |
| --- | --- | --- | --- |
| **Sex** |  |  |  |
| Female | 19 (51%) | 40 (46%) | 3 (60%) |
| Male | 18 (49%) | 47 (54%) | 2 (40%) |
| **Age categories** |  |  |  |
| 0-9 years old | 1 (3%) | 15 (17%) | 1 (20%) |
| 10-19 years old | 2 (5%) | 23 (27%) | 0 |
| 20-29 years old | 4 (11%) | 6 (7%) | 0 |
| 30-39 years old | 9 (24%) | 11 (13%) | 4 (80%) |
| 40-49 years old | 15 (41%) | 16 (18%) | 0 |
| ≥50 years old | 6 (16%) | 16 (18%) | 0 |
| **Pre-existing conditions** |  |  |  |
| None | 30 (81%) | 67 (77%) | 4 (80%) |
| At least one | 7 (19%) | 20 (23%) | 1 (20%) |
| • Obesity | 4 (11%) | 9 (10%) | 0 |
| • Cancer | 0 | 1 (1%) | 0 |
| • Chronic heart condition | 0 | 2 (2%) | 0 |
| • Asthma | 1 (3%) | 4 (5%) | 0 |
| • Chronic lung condition | 0 | 1 (1%) | 0 |
| • Chronic hematologic condition | 0 | 1 (1%) | 0 |
| • Chronic neurologic condition | 3 (8%) | 3 (3%) | 1 (20%) |
| • Other condition | 4 (11%) | 7 (8%) | 0 |
| **Reported symptoms** |  |  |  |
| None | 0 | 24 (28%) | 0 |
| At least one | 37 (100%) | 63 (72%) | 5 (100%) |
| • Fever | 23 (62%) | 22 (25%) | 4 (80%) |
| • Sore throat | 19 (51%) | 31 (35%) | 2 (40%) |
| • Cough | 28 (76%) | 35 (40%) | 4 (80%) |
| • Runny nose | 13 (35%) | 32 (37%) | 5 (100%) |
| • Shortness of breath | 12 (32%) | 14 (16%) | 3 (60%) |
| • Chills | 26 (72%) | 23 (27%) | 5 (100%) |
| • Vomiting | 3 (9%) | 3 (4%) | 0 |
| • Nausea | 10 (29%) | 16 (20%) | 0 |
| • Diarrhea | 12 (35%) | 20 (24%) | 2 (40%) |
| • Headache | 24 (73%) | 37 (45%) | 2 (40%) |
| • Rash | 3 (10%) | 3 (4%) | 0 |
| • Conjunctivitis | 0 | 2 (2%) | 0 |
| • Myalgia | 27 (77%) | 23 (28%) | 2 (40%) |
| • Arthralgia | 16 (48%) | 17 (21%) | 2 (40%) |
| • Loss of appetite | 20 (57%) | 17 (20%) | 0 |
| • Nose bleed | 3 (9%) | 7 (8%) | 3 (75%) |
| • Fatigue | 35 (95%) | 38 (44%) | 4 (80%) |
| • General malaise | 20 (56%) | 15 (18%) | 1 (20%) |
| • Altered consciousness | 2 (6%) | 3 (4%) | 0 |
| • Anosmia | 22 (63%) | 18 (22%) | 4 (80%) |

### Household transmission

#### Participants

In six households, one of the other COVID-19 cases had had earlier onset of the symptoms than the index case, which led to reclassification to another household member as primary case and of index cases as secondary cases. In two households, at least one secondary case had the same timing of onset as the index case, which led to exclusion of five participants from two households. Hence, we included 124 participants from 37 different households in the household transmission component of the study.

Due to restricted access to RT-PCR testing for SARS-CoV-2 and delayed availability of test results during the early epidemic, households were recruited, on average, 28 days [median, IQR: 21-40] after symptom onset of the index case. Hence only four households were visited around day 7, day 14 and day 28; 13 households were visited around day 14 and day 28, while in 20 households, only one visit was conducted. On average, the last household transmission study visit was conducted after median of 39 days [IQR: 35-43] since onset of the primary case (Figure 1).

**Figure 1:**
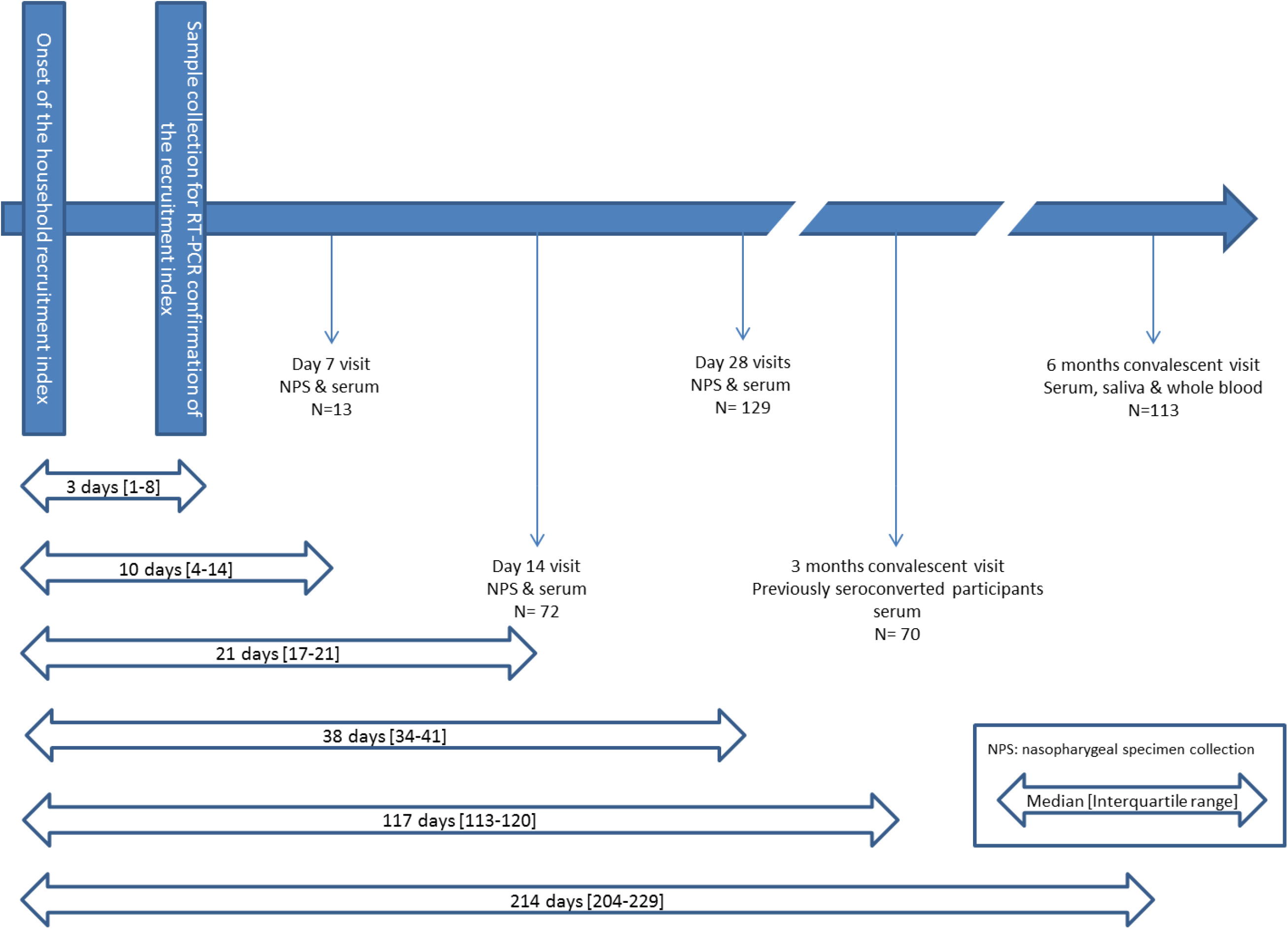
SARS-CoV-2 household transmission, Finland 2020: study design and timeline of recruitment and follow-up (N total, 129 participants)

#### Household transmission dynamics

Following SARS-CoV-2 infection of primary cases, 42 out of 87 household members developed an infection confirmed either via RT-PCR (n=28, 32%) or serology (n=42, 48%) by the end of household transmission follow-up. Seropositivity was defined by detection of IgG Ab to SARS-CoV-2 nucleoprotein (n=42) and/or NAb (n=40). In 13/37 households, no secondary cases were identified. Overall, the SAR was 48% [IC_95%_: 38%-59%]; 50% [IQR: 0-100%] on a household level, with a median effective reproduction number of 1 [IQR: 0-2] (Table 2)

**Table 2:** Household transmission dynamics.

|  | Secondary cases<br>(N=42) | Household contacts<br>(N=87) | Percentage of<br>secondary cases | Effective<br>reproduction<br>number |
| --- | --- | --- | --- | --- |
|  | N | N | % | Median [IQR] |
| Age of the primary case |  |  |  |  |
| 0-9 years old (n=1) | 0 | 3 | 0% | 0 |
| 10-19 years old (n=2) | 7 | 7 | 100% | 3.5 [3-4] |
| 20-29 years old (n=4) | 7 | 13 | 54% | 2 [1.5-2] |
| 30-39 years old | 6 | 14 | 43% | 1 [0-1] |
| 40-49 years old | 16 | 41 | 39% | 1 [0-2] |
| ≥50 years old (n=6) | 6 | 9 | 67% | 1 [0-2] |
| Household size |  |  |  |  |
| 2 members (n=11) | 7 | 11 | 64% | 1 [0-1] |
| 3 members (n=8) | 6 | 16 | 38% | 0 [0-2] |
| 4 members (n=13) | 17 | 39 | 44% | 1 [1-2] |
| 5 members (n=4) | 7 | 16 | 44% | 1.5 [0.5-3] |
| 6 members (n=1) | 5 | 5 | 100% | 5 [5-5] |
| Rooms |  |  |  |  |
| <1 room per household member | 24 | 52 | 46% | 1 [0-2] |
| 1-2 rooms per household members | 16 | 27 | 59% | 1 [1-2] |
| >2 rooms per household members | 2 | 8 | 25% | 0 [0-1] |
| Bedrooms |  |  |  |  |
| <1 bedroom per household member | 37 | 75 | 49% | 1 [0-2] |
| At least one bedroom per household members | 5 | 12 | 42% | 0.5 [0-1] |
| Total | 42 | 87 | 48% | 1 [IQR: 0-2] |

#### Description of secondary cases and healthy household contacts

We did not find any significant differences regarding age, sex, type of relationship to the primary case and previous comorbidities between secondary cases and household contacts who did not get infected. Out of 42 secondary cases, 88% [IC_95%_: 74%-95%] (n=37) reported at least one symptom, while 58% [IC_95%_: 43%-72%] (26/45) of healthy household contacts reported they had experienced symptoms between primary case onset and the end of follow-up (p-value=0.002). The most reported symptoms among secondary cases were cough (n=27/42, 64%) and headache (n=26/39, 67%) (Table 3).

**Table 3:** Secondary cases and healthy household contacts characteristics and symptoms.

|  | Healthy household contacts<br>(N=45) | Secondary cases<br>(N=42) | p-value |
| --- | --- | --- | --- |
|  | N (%) | N (%) |  |
| **Sex** |  |  |  |
| Female | 19 (42%) | 21 (50%) | 0.47 |
| Male | 26 (58%) | 21 (50%) |  |
| **Age groups** |  |  |  |
| 0-9 years old | 10 (22%) | 5 (12%) | 0.86 |
| 10-19 years old | 11 (24%) | 12 (29%) |  |
| 20-29 years old | 3 (7%) | 3 (7%) |  |
| 30-39 years old | 6 (13%) | 5 (12%) |  |
| 40-49 years old | 7 (16%) | 9 (21%) |  |
| ≥50 years old | 8 (18%) | 8 (19%) |  |
| **Relationship to the primary case** |  |  |  |
| Child | 21 (47%) | 13 (31%) | 0.46 |
| Parent/Step-parent | 9 (20%) | 9 (21%) |  |
| Spouse/partner | 12 (27%) | 15 (36%) |  |
| Sibling | 3 (7%) | 5 (12%) |  |
| **Diagnosis method** |  |  |  |
| Positive PCR-testing | 0 | 28 (67%) |  |
| Positive MNT-testing | 0 | 40 (95%) |  |
| Positive FMIA testing | 0 | 42 (100%) |  |
| **Pre-existing conditions** |  |  |  |
| None | 36 (80%) | 31 (74%) | 0.49 |
| At least one | 9 (20%) | 11 (26%) |  |
| • Obesity | 4 (9%) | 5 (12%) | 0.67 |
| • Cancer | 1 (2%) | 0 | 1.00 |
| • Chronic heart condition | 0 | 2 (5%) | 0.23 |
| • Asthma | 3 (7%) | 1 (2%) | 0.62 |
| • Chronic lung condition | 0 | 1 (2%) | 0.48 |
| • Chronic hematologic condition | 1 (2%) | 0 | 1.00 |
| • Chronic neurologic condition | 2 (4%) | 1 (2%) | 1.00 |
| • Other conditions | 1 (2%) | 6 (14%) | 0.11 |
| **Reported symptoms** |  |  |  |
| None | 19 (42%) | 5 (12%) | 0.002 |
| At least one | 26 (58%) | 37 (88%) |  |
| • Fever | 2 (4%) | 20 (48%) | 0.000 |
| • Sore throat | 11 (24%) | 20 (48%) | 0.024 |
| • Cough | 8 (18%) | 27 (64%) | 0.000 |
| • Runny nose | 11 (24%) | 21 (50%) | 0.014 |
| • Shortness of breath | 3 (7%) | 11 (26%) | 0.013 |
| • Chills | 6 (13%) | 17 (44%) | 0.002 |
| • Vomiting | 1 (2%) | 2 (5%) | 0.488 |
| • Nausea | 5 (11%) | 11 (29%) | 0.045 |
| • Diarrhea | 6 (14%) | 14 (35%) | 0.022 |
| • Headache | 11 (26%) | 26 (67%) | 0.000 |
| • Rash | 1 (2%) | 2 (5%) | 0.599 |
| • Conjunctivitis | 0 | 2 (5%) | 0.218 |
| • Myalgia | 4 (9%) | 19 (50%) | 0.000 |
| • Arthralgia | 4 (9%) | 13 (33%) | 0.007 |
| • Loss of appetite | 3 (7%) | 14 (34%) | 0.002 |
| • Nose bleed | 3 (7%) | 4 (10%) | 0.702 |
| • Fatigue | 6 (14%) | 14 (35%) | 0.022 |
| • General malaise | 6 (14%) | 9 (23%) | 0.286 |
| • Altered consciousness | 1 (2%) | 2 (5%) | 0.595 |
| • Anosmia | 0 | 18 (46%) | 0.000 |

### Development and follow-up of SARS-CoV-2 immunity

In total, 80 (62%) participants had developed both SARS-CoV-2 IgG Ab and NAb including 63 out of 64 participants with documented RT-PCR confirmation of SARS-CoV-2 infection. Three participants had only developed IgG Ab, none of whom with a RT-PCR documented infection, by the end of the household transmission study follow-up (Table 4).

**Table 4:** SARS-CoV-2 confirmed cases: immunity status over time.

|  | Initial follow up | 3 months convalescent visit | 6 months convalescent visit |
| --- | --- | --- | --- |
|  | N (%) | N (%) | N (%) |
| IgG antibodies to SARS-CoV-2 nucleoprotein detection |  |  |  |
| • Positive | 83 (99%) | 68 (99%) | 59 (78%) |
| • Negative | 1 (1%) | 1 (1%) | 17 (22%) |
| SARS-CoV-2 neutralizing antibodies testing |  |  |  |
| • Positive | 80 (95%) | 63 (90%) | 63 (83%) |
| • Negative | 4 (5%) | 3 (4%) | 12 (16%) |
| • Inconclusive or borderline | 0 | 4 (6%) | 1 (1%) |

#### Follow-up of immunity at three months

Out of 80 participants positive for SARS-CoV-2 IgG Ab and NAb, 70 (87.5%) participated in 3-month sample collection at median 88 days [IQR: 81-99] after first sample collection. Out of 69 participants tested by both methods, 62 (90%, [IC_95%_: 80%-95%]) still had both IgG Ab and NAb, while 6 participants (9% [IC_95%_:4%-18%]) only had detectable IgG Ab. One participant was tested positive for NAb only due to lack of specimen volume.

#### Follow-up of immunity at six months

Out of 129 participants from the initial follow-up, 113 (88%) underwent the 6-month convalescent visit and 111 serum samples were collected at a median of 185 [IQR: 178-198] days after the first sample collection. Out of 76 cases identified during the initial follow-up, 59 (78%, [IC_95%_: 67%-86%]) and 63 (83%, [IC_95%_: 73%-90%]) still had detectable IgG Ab to SARS-CoV-2 nucleoprotein and NAb, respectively (Table 4). Additionally, we observed seroconversion in two household contacts between the initial follow-up and the 6-month convalescent visit: One participant now had detectable IgG Ab to SARS-CoV-2 nucleoprotein without knowledge of exposure to a confirmed case, a symptomatic contact or reporting any symptoms, while another household contact now had both NAb and detectable IgG Ab and reported symptoms, both for themselves and in their close circle.

#### Trends of antibody levels in confirmed cases

##### IgG antibodies to SARS-CoV-2 nucleoprotein

In the samples collected from confirmed cases during the initial follow-up of the household transmission study, we observed a progressive increase in the mean concentration of IgG Ab to SARS-CoV-2 nucleoprotein peaking at 1 month post onset of the index case: 32.72 U/mL (GMC, IC95%: 23.63-45.30) (Figure 2). In average, the highest concentration of IgG Ab to SARS-CoV-2 nucleoprotein was 35.65 (GMC, IC95%: 26.04-48.81) during the household transmission study, followed by a subsequent decline to 15.12 U/mL (GMC, IC95%: 11.45-19.98) and 3.33 U/mL (GMC, IC95%: 2.45-4.53) at the 3 and 6 months convalescent visits, respectively (Table 5). Across all time points, SARS-CoV-2 IgG Ab concentrations were higher among participants who had required hospital care compared to cases with mild symptoms. Compared to the highest IgG Ab concentration during the initial follow-up, there was a 66% (median, IQR: 48% - 81%) decrease in 66/69 participants and a 53% (median, IQR : 17% - 257%) increase in three participants at three months. At six months, IgG Ab concentration had decreased by 89% (median, IQR: 81% - 95%) in all confirmed cases.

**Table 5:** Trends in mean IgG antibody concentration (GMC) to SARS-CoV-2 nucleoprotein.

|  | Initial follow-up<sup>§</sup> | 3 months convalescent visit | 6 months convalescent visit |
| --- | --- | --- | --- |
|  | GMC [IC<sub>95%</sub>] | GMC [IC<sub>95%</sub>] | GMC [IC<sub>95%</sub>] |
| All confirmed cases* | N=84<br>35.65[26.03-48.81] | N=69<br>15.12 [11.45-19.98] | N=76<br>3.33 [2.45-4.53] |
| • Asymptomatic cases | N=5<br>22.67[2.22-231.86] | N=3<br>20.31 [0.31-1339.19] | N=3<br>5.11 [0.33-78.86] |
| • Cases with mild symptoms | N=75<br>33.70 [24.35-46.64] | N=62<br>13.57 [10.27-17.93] | N=69<br>3.02 [2.18-4.17] |
| • Cases requiring hospital care | N=4<br>179.79 [85.13-379.73] | N=4<br>64.79 [14.02-299.33] | N=4<br>13.11 [5.56-30.90] |

<sup>§</sup> highest value obtained during initial follow-up
\*Confirmed cases as defined during initial follow-up

**Figure 2:**
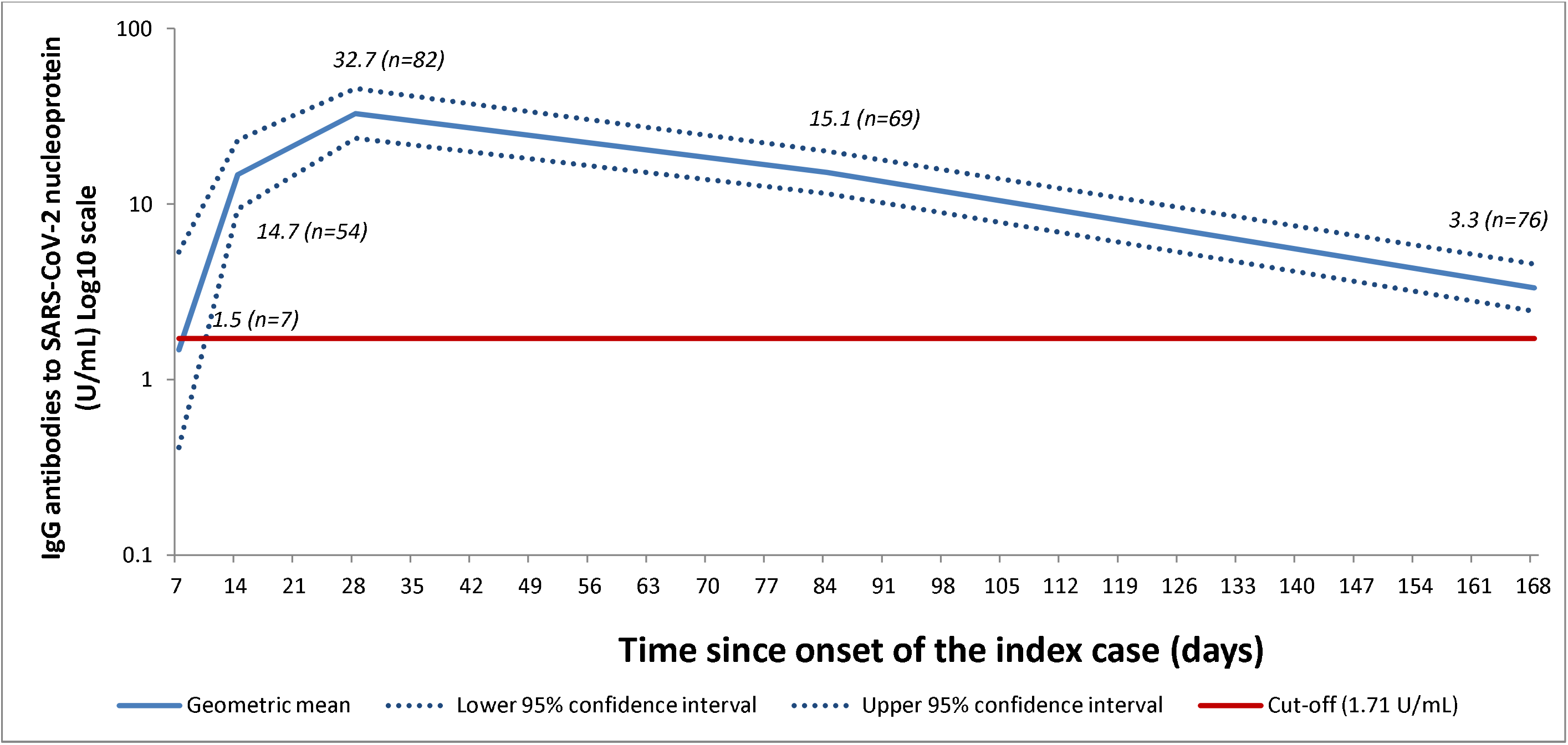
Development and follow-up of IgG antibodies to SARS-CoV-2 nucleoprotein among confirmed cases, geometric mean and 95% confidence interval.

##### SARS-Cov-2 neutralizing antibodies

As evidenced with IgG Ab, NAb titers progressively increased until the final visit at 28 days both for Fin1-20, peaking at 39.14 (GMT, IC95%: 29.95-51.17) (Figure 3) and Fin25-20 strain (not shown). We re-analysed the last available specimen from the initial follow up in the same assay as the ones collected during the convalescent visits at three and six months for each previously confirmed case. NAb titers decreased from 40.27 (GMT, IC95%: 30.16-53.76) to 19.58 (GMT, IC95%: 15.05-25.49) and 16.89 (GMT, IC95%: 12.77-22.35) at the three and six months convalescent visits, respectively, for the Fin1-20 strain (Table 6). While for the Fin25-20 strain, we observed a NAb titer of 33.46 (GMT, IC95%: 24.88-44.99) at the 28 days visit, decreasing to 13.26 (GMT, IC95%: 10.01-17.55) at three months and 11.75 (GMT, IC95%: 9.10-15.17) at 6 months. As evidenced with IgG Ab, SARS-CoV-2 NAb titers were higher among participants who had required hospital care compared to cases with mild symptoms across all time points (Table 6).

**Table 6:** Trends in mean neutralizing antibody titers (GMT) to SARS-CoV-2.

|  | 28 days visit<sup>§</sup> | 3 months convalescent visit | 6 months convalescent visit |
| --- | --- | --- | --- |
|  | GMT [IC<sub>95%</sub>] | GMT [IC<sub>95%</sub>] | GMT [IC<sub>95%</sub>] |
| **Fin1-20 neutralizing antibodies titers** |  |  |  |
| All confirmed cases* | N=68 | N=61 | N=75 |
|  | 40.27 [30.16-53.76] | 19.59 [15.05-25.49] | 16.89 [12.77-22.35] |
| • Asymptomatic cases | N=3 | N=3 | N=3 |
|  | 46.16 [9.55-223.09] | 20.16 [7.46-54.48] | 16.64 [1.80-153.75] |
| • Cases with mild symptoms | N=61 | N=54 | N=68 |
|  | 34.57 [26.05-45.87] | 17.51 [13.34-22.98] | 15.70 [11.68-21.09] |
| • Cases requiring hospital care | N=4 | N=4 | N=4 |
|  | 372.86 [236.91-586.82] | 86.75 [33.33-225.78] | 59.56 [19.92-178.11] |
| **Fin25-20 neutralizing antibodies titers** |  |  |  |
| All confirmed cases* | N=67 | N=60 | N=74 |
|  | 33.46 [24.88-44.99] | 13.26 [10.01-17.55] | 11.75 [9.10-15.17] |
| • Asymptomatic cases | N=3 | N=3 | N=3 |
|  | 33.28 [14.01-79.05] | 11.54 [4.86-27.40] | 12.00 [2.14-67.14] |
| • Cases with mild symptoms | N=60 | N=53 | N=67 |
|  | 28.83 [21.55-38.56] | 11.90 [8.89-15.94] | 10.81 [8.30-14.09] |
| • Cases requiring hospital care | N=4 | N=4 | N=4 |
|  | 313.53 [127.34-772.01] | 61.34 [25.85-145.56] | 46.61 [13.74-158.04] |

<sup>§</sup> If 28 days visit samples were not available for re-analysis, samples from previous visit was used
\*Confirmed cases as defined during initial follow-up

**Figure 3:**
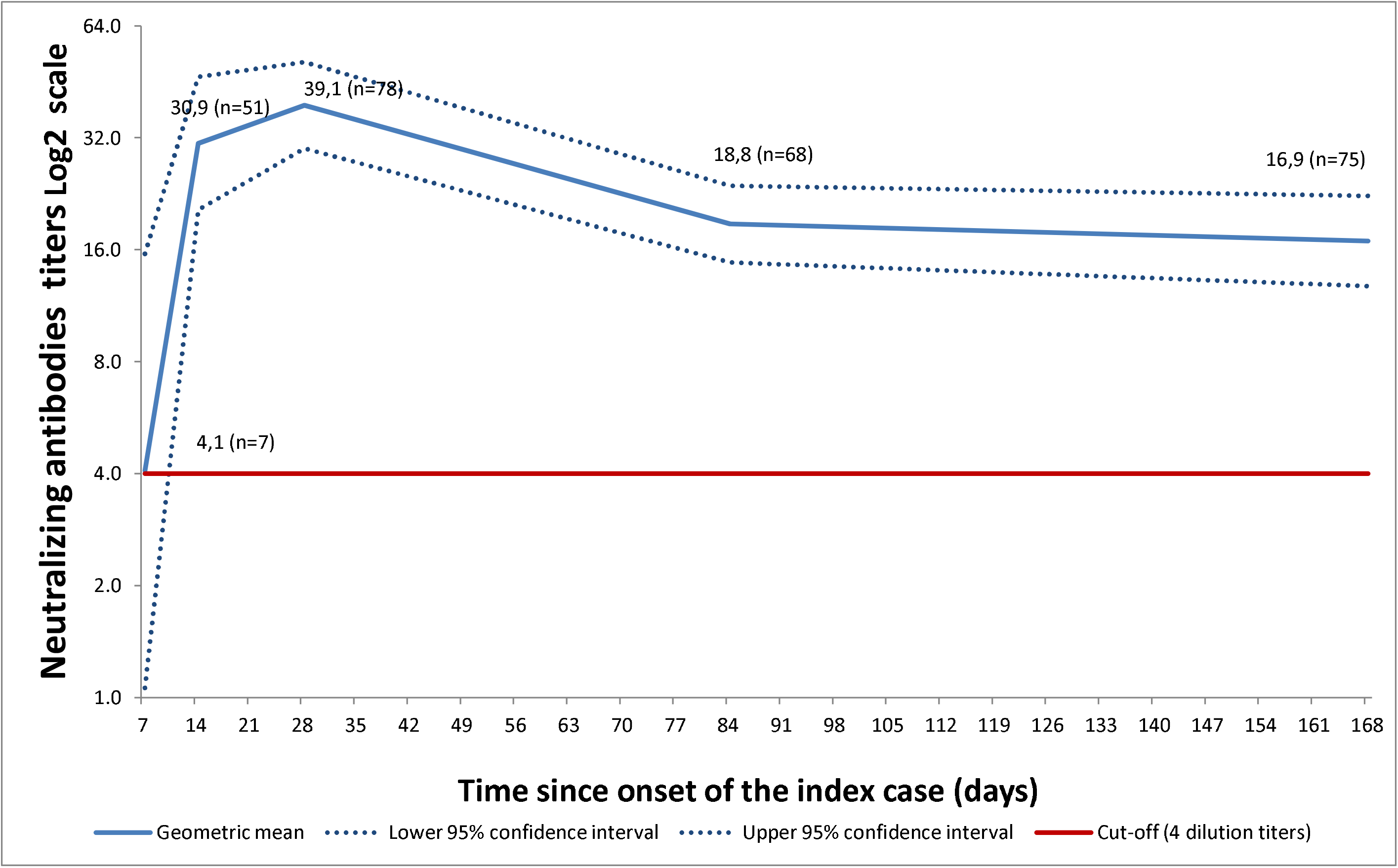
Trends in neutralizing antibody titers over time: Fin1-20.

On an individual level, compared to the last sample available from the initial household transmission follow-up, the Fin1-20 strain GMT NAb titers decreased by 62.5% (median, IQR:50%-75%) for 49 participants, remained the same for 4 participants and increased by 100% (median, IQR:100%-100%) for 6 participants at three months. At six months, the Fin1-20 strain NAb titers decreased by 67% (median, IQR:59%-81%) for 52 participants, remained the same for 8 participants and increased by 167% (median, IQR:75%-200%) for 8 participants, compared to the initial household transmission follow-up.

## Discussion

Exposure to an individual with SARS-CoV-2 in a household led to secondary infection in half of the household contacts (SAR 48%). Over the follow-up period, only 5 (12%) individuals with confirmed SARS-CoV-2 infection reported experiencing no symptoms. Convalescent sampling of patients showed that at 6 months after infection, SARS-CoV-2 NAb and IgG Ab to SARS-CoV-2 nucleoprotein were still detected in the large majority of the initially seropositive participants. There was only a very small decline in the IgG Ab and NAb concentrations from 3 months to 6 months, suggesting that the antibody levels have reached a plateau where they may persist for a longer period. We will further assess the long-term persistence of immunity in a follow-up study of the household cohort, where IgG Ab and NAb will be measured from samples collected at 12 months following infection.

Depending on the duration of follow-up and the diagnostic methods, the estimates obtained by previous SARS-CoV-2 household transmission studies are very heterogeneous, with SARs ranging from 4.6% to 89.8%.^13–26^ However our findings were in line with other studies where seroconversion was used for identification of secondary cases over a long duration of follow-up. In the Netherlands, using RT-PCR testing and SARS-CoV-2 IgG enzyme-linked immunosorbent assay (ELISA), Reukers et al. estimated that the overall household SAR was 43% (95%CI 33%-53%) after 4-6 weeks of follow-up.^27^ While in the United States, when using a CDC-developed SARS-CoV-2 ELISA, Lewis et al. showed that after 14 days of follow-up, the SAR was 29% (95%CI: 23–36%) overall.^28^ It is plausible that our estimate of the SAR, as ones from previously published studies are underestimated, knowing that not all who are infected will develop detectable antibodies but could still develop T-cell responses^29^.

Regarding long-term persistence of antibodies following COVID-19 infection, published data remains scarce. In the Netherlands, the study of a nationwide sample showed that 6 months after initial seroconversion, 99% of patients still had detectable anti-SARS-CoV-2 Spike S1 IgG antibodies, while in Bengbu (China), 95.5% (n=21) of convalescent patients were still IgG positive in a chemiluminescence immunoassay using recombinant antigens based on the SARS-CoV-2 spike protein at 6 months post symptom onset^30,31^. In Italy, Dittadi et al. analyzed 20 specimens at 85 to 187 days post-onset of COVID-19 and identified a 80% and 100% positivity rate using commercial assays based on the S1, S2 and N proteins.^32^ In Belgium, the follow-up of a cohort of healthcare workers showed that out of 81 IgG positive healthcare workers tested monthly with a commercial semi-quantitative ELISA (Euroimmun IgG; Medizinische Labordiagnostika, Lübeck, Germany) targeting SARS-CoV-2 S1, 74 remained positive including 67 healthcare workers who had had detectable antibodies for 120 days or more.^33^ Similar findings were also reported by Isho et al. at 3 months post-symptom onset in a cross-sectional survey, as well as by Wang et al. who followed 113 previously hospitalized patients during a close to three months convalescent phase.^34,35^ Finally, as far as IgG Ab persistence is concerned, the follow-up of previously positive HCWs in Eastern France showed a 20.1% positivity rate of IgG Ab to SARS-CoV-2 nucleoprotein at 11-13 months post onset^36^.

Results from studies where NAb were investigated at 3 or more months post symptom onset varied widely, which might also be the consequence of the different neutralization test methods and starting serum dilutions being used. One conducted in Sri-Lanka showed that SARS-CoV-2 NAb titers were higher in patients presenting with a more severe disease compared to cases with milder symptoms. This is in line with our findings, even though, in their study no NAb were detected in 50% of convalescent patients at 90 days.^37^ In China, Long et al. observed that at 8 weeks after hospital discharge, patients who had had a symptomatic disease had a slightly lower relative decrease of neutralizing serum antibody levels (8.3% in median) compared to asymptomatic cases (11.7% in median) with more asymptomatic individuals becoming seronegative.^38^ In a cohort of 26 French healthcare workers, 85% of them still had NAb at 3 months after disease onset, while in Switzerland, it was estimated that 93% of 196 convalescent healthcare workers still had NAb at 6 months post infection.^39,40^ Additionally, when investigating the decline of NAb titers at 5 to 6 months after disease onset, Wajnberg et al. and Pradenas et al. identified that NAb titers remained relatively stable despite a mild decrease in patients with mild symptoms or requiring hospitalization.^41,42^

Our study suggests long term persistence of antibodies following infection. However, as no correlate of protection has been established for SARS-CoV-2 infection, it remains unknown what level of NAb titer would provide protection against reinfection or disease.

Our study had two main limitations: First, this was not a randomly selected cohort of households as we recruited participants through a voluntary convenience sampling. Second, due to the lag between onset and availability of RT-PCR results, most participants were recruited retrospectively, which prevented us from conducting early household visits to estimate serial intervals for all subjects. Additionally, our estimation of the effective reproductive number was influenced by the limited size of households, with median 3 [IQR:2-4] members. However, this was in line with the demographics of Helsinki, where only 51.5% of households consisted of at least two members including 20.8% of households with 3 or more members.^43^ Finally, our limited sample size did not allow for comparison of secondary transmission rate depending on age-group of the primary case.

Our main strengths were that, unlike most household transmission studies, we measured not only IgG Ab but also NAb to SARS-CoV-2, and that household transmission was assessed over a one month period, providing enough time for the development of antibodies, in case of secondary infection. Finally, only a few participants were lost to follow-up, as 113/129 participated to a convalescent visit at 6 months, allowing us to assess trends in IgG Ab and NAb for 111 study participants out of 129 over a long time period.

When living in the same household of an RT-PCR confirmed case, the risk of transmission for other household members is extremely high as 1 out of 2 get infected. The challenge is that this is a setting in which adherence to infection prevention and control measures is conceivably low. Household members at risk of severe COVID-19 should be informed of this risk and consideration should be given to the isolation of the cases in separate facilities, even though viral transmission might have already occurred in other household members before diagnosis^44^.

## Data Availability

Individual data remains the sole property of the Finnish institute for Health and Welfare and will not be shared. Additional analyses can be requested to the corresponding author.

## Authors’ contributions

TD, HN, NI, MM and AAP conceived and designed the study. TL supervised contact tracing as well as helped in the recruitment process. TD, LH, OL and HN recruited participants to the study and collected samples and data. NI developed, supervised and interpreted the virological analysis, PÖ cultivated the viral strains for immunological analyses. CV, AH, AS, OL, SV, KL, HV, HH and MM developed, performed, supervised and interpreted the immunological analyses. TD performed the statistical analyses andwrote the first version of the report. All authors critically reviewed and approved the final version of the report

## Acknowledgements

We would like to thank all study participants for joining our research. We thank Marja-Liisa Ollonen, Tiina Pulliainen, Raisa Hanninen, Mervi Nosa, Leena Saarinen Marja Suorsa, Esa Ruokokoski, Henna Makela, Veronica Cristea, Juha Oksanen and Anna-Leena Lohiniva from the Finnish institute for Health and Welfare for their expert technical assistance and support in conducting this study. We also wish to thank the World Health Organisation Unity studies initiative with Isabel Bergeri for her input on our protocol and Rebecca Grant for her input on our protocol and thorough review of our manuscript. Finally, we gratefully acknowledge the authors and their respective laboratories, who analyzed and submitted the sequences to GISAID’s EpiCoVTM Database

## References

1 WHO | Pneumonia of unknown cause – China. https://www.who.int/csr/don/05-january-2020-pneumonia-of-unkown-cause-china/en/ãfbclid=IwAR2v89e9lp70O6GTra13FIPHCLw4WJ8kL20Uylx5zZNtWAYvbR0sEATr_rg (accessed Dec 6, 2020).

2 WHO Director-General’s opening remarks at the media briefing on COVID-19 - 11 March 2020. https://www.who.int/director-general/speeches/detail/who-director-general-s-opening-remarks-at-the-media-briefing-on-covid-1911-march-2020 (accessed Dec 6, 2020).

3 WHO Coronavirus Disease (COVID-19) Dashboard. https://covid19.who.int (accessed Dec 6, 2020).

4 Cristea V, Dub T, Luomala O, Sivelä J. COVID-19 behavioural insights study: Preliminary findings from Finland, April-May, 2020. medRxiv 2020; : 2020.10.11.20210724.

5 Household transmission investigation protocol for 2019-novel coronavirus (COVID-19) infection. https://www.who.int/publications-detail-redirect/household-transmission-investigation-protocol-for-2019-novel-coronavirus-(2019-ncov)-infection (accessed July 2, 2020).

6 COVID-19 Coronavirus infection and exposure in households THL. Finn. Inst. Health Welf. THL Finl. https://thl.fi/en/web/thlfi-en/research-and-development/research-and-projects/covid-19-coronavirus-infection-and-exposure-in-households (accessed May 16, 2021).

7 Corman VM, Landt O, Kaiser M, et al. Detection of 2019 novel coronavirus (2019-nCoV) by real-time RT-PCR. Eurosurveillance 2020; 25. DOI:10.2807/1560-7917.ES.2020.25.3.2000045.

8 Ekström N, Virta C, Haveri A, et al. Analytical and clinical evaluation of antibody tests for SARS-CoV-2 serosurveillance studies used in Finland in 2020. medRxiv 2021; : 2021.01.21.21250207.

9 Haveri A, Smura T, Kuivanen S, et al. Serological and molecular findings during SARS-CoV-2 infection: the first case study in Finland, January to February 2020. Eurosurveillance 2020; 25 2000266.

10 National Institute for Biological Standards and Control. First WHO International Standard for anti SARS-CoV-2 immunoglobulin (human) NIBSC code: 20/136. 2020; published online Dec 17. https://www.nibsc.org/documents/ifu/20-136.pdf.

11 Ajantasainen lainsäädäntö: Laki Terveyden ja hyvinvoinnin laitoksesta 668/2008. https://www.finlex.fi/fi/laki/ajantasa/2008/20080668 (accessed July 12, 2020).

12 Ajantasainen lainsäädäntö: Tartuntatautilaki 1227/2016. https://www.finlex.fi/fi/laki/ajantasa/2016/20161227 (accessed July 12, 2020).

13 Fung HF, Martinez L, Alarid-Escudero F, et al. The household secondary attack rate of SARS-CoV-2: A rapid review. Clin Infect Dis Off Publ Infect Dis Soc Am 2020; published online Oct 12. DOI:10.1093/cid/ciaa1558.

14 Grijalva CG, Rolfes MA, Zhu Y, et al. Transmission of SARS-COV-2 Infections in Households Tennessee and Wisconsin, April-September 2020. MMWR Morb Mortal Wkly Rep 2020; 69: 1631–4.

15 Horchinbilig U, Gao Y, Chang H, et al. Investigation of 100 SARS-CoV-2 infected families in Wuhan: Transmission patterns and follow-up. J Glob Health 2020; 10: 021103.

16 Lei H, Xu X, Xiao S, Wu X, Shu Y. Household transmission of COVID-19-a systematic review and meta-analysis. J Infect 2020; 81: 979–97.

17 Madewell ZJ, Yang Y, Longini IM, Halloran ME, Dean NE. Household Transmission of SARS-CoV-2: A Systematic Review and Meta-analysis. JAMA Netw Open 2020; 3: e2031756.

18 Shah K, Kandre Y, Mavalankar D. Secondary attack rate in household contacts of COVID-19 Paediatric index cases: a study from Western India. J Public Health Oxf Engl 2021; published online Jan 18. DOI:10.1093/pubmed/fdaa269.

19 Angulo-Bazán Y, Solis-Sánchez G, Cardenas F, Jorge A, Acosta J, Cabezas C. Household transmission of SARS-CoV-2 (COVID-19) in Lima, Peru. Cad Saude Publica 2021; 37: e00238720.

20 Cerami C, Rapp T, Lin F-C, et al. High household transmission of SARS-CoV-2 in the United States: living density, viral load, and disproportionate impact on communities of color. medRxiv 2021; published online March 12. DOI:10.1101/2021.03.10.21253173.

21 Galow L, Haag L, Kahre E, et al. Lower household transmission rates of SARS-CoV-2 from children compared to adults. J Infect 2021; published online April 28. DOI:10.1016/j.jinf.2021.04.022.

22 Gomaa MR, El Rifay AS, Shehata M, et al. Incidence, household transmission, and neutralizing antibody seroprevalence of Coronavirus Disease 2019 in Egypt: Results of a community-based cohort. PLoS Pathog 2021; 17: e1009413.

23 Kuba Y, Shingaki A, Nidaira M, et al. The characteristics of household transmission during COVID-19 outbreak in Okinawa, Japan from February to May 2020. Jpn J Infect Dis 2021; published online April 30. DOI:10.7883/yoken.JJID.2020.943.

24 Kuwelker K, Zhou F, Blomberg B, et al. Attack rates amongst household members of outpatients with confirmed COVID-19 in Bergen, Norway: A case-ascertained study. Lancet Reg Health Eur 2021; 3: 100014.

25 Reukers DFM, van Boven M, Meijer A, et al. High infection secondary attack rates of SARS-CoV-2 in Dutch households revealed by dense sampling. Clin Infect Dis Off Publ Infect Dis Soc Am 2021; published online April 2. DOI:10.1093/cid/ciab237.

26 Salihefendic N, Zildzic M, Huseinagic H, Ahmetagic S, Salihefendic D, Masic I. Intrafamilial Spread of COVID-19 Infection Within Population in Bosnia and Herzegovina. Mater Socio-Medica 2021; 33: 4–9.

27 Reukers DFM, van Boven M, Meijer A, et al. High infection attack rates of SARS-CoV-2 in Dutch households revealed by dense sampling. medRxiv 2021; : 2021.01.26.21250512.

28 Lewis NM, Chu VT, Ye D, et al. Household Transmission of SARS-CoV-2 in the United States. Clin Infect Dis Off Publ Infect Dis Soc Am 2020; published online Aug 16. DOI:10.1093/cid/ciaa1166.

29 Fafi-Kremer S, Bruel T, Madec Y, et al. Serologic responses to SARS-CoV-2 infection among hospital staff with mild disease in eastern France. Infectious Diseases (except HIV/AIDS), 2020 DOI:10.1101/2020.05.19.20101832.

30 den Hartog G, Vos ERA, van den Hoogen LL, et al. Persistence of antibodies to SARS-CoV-2 in relation to symptoms in a nationwide prospective study. Clin Infect Dis Off Publ Infect Dis Soc Am 2021; published online Feb 24. DOI:10.1093/cid/ciab172.

31 Liu C, Yu X, Gao C, et al. Characterization of antibody responses to SARS-CoV-2 in convalescent COVID-19 patients. J Med Virol 2021; 93: 2227–33.

32 Dittadi R, Afshar H, Carraro P. Two SARS-CoV-2 IgG immunoassays comparison and time-course profile of antibodies response. Diagn Microbiol Infect Dis 2020; 99: 115297.

33 Duysburgh E, Mortgat L, Barbezange C, et al. Persistence of IgG response to SARS-CoV-2. Lancet Infect Dis 2021; 21: 163–4.

34 Isho B, Abe KT, Zuo M, et al. Persistence of serum and saliva antibody responses to SARS-CoV-2 spike antigens in COVID-19 patients. Sci Immunol 2020; 5. DOI:10.1126/sciimmunol.abe5511.

35 Wang Y, Li J, Li H, Lei P, Shen G, Yang C. Persistence of SARS-CoV-2-specific antibodies in COVID-19 patients. Int Immunopharmacol 2021; 90: 107271.

36 Gallais F, Gantner P, Bruel T, et al. Anti-SARS-CoV-2 Antibodies Persist for up to 13 Months and Reduce Risk of Reinfection. medRxiv 2021; : 2021.05.07.21256823.

37 Jeewandara C, Jayathilaka D, Gomes L, et al. SARS-CoV-2 neutralizing antibodies in patients with varying severity of acute COVID-19 illness. Sci Rep 2021; 11: 2062.

38 Long Q-X, Tang X-J, Shi Q-L, et al. Clinical and immunological assessment of asymptomatic SARS-CoV-2 infections. Nat Med 2020; 26: 1200–4.

39 L’Huillier AG, Meyer B, Andrey DO, et al. Antibody persistence in the first 6 months following SARS-CoV-2 infection among hospital workers: a prospective longitudinal study. Clin Microbiol Infect Off Publ Eur Soc Clin Microbiol Infect Dis 2021; published online Jan 20. DOI:10.1016/j.cmi.2021.01.005.

40 Marot S, Malet I, Leducq V, et al. Rapid decline of neutralizing antibodies against SARS-CoV-2 among infected healthcare workers. Nat Commun 2021; 12: 844.

41 Wajnberg A, Amanat F, Firpo A, et al. SARS-CoV-2 infection induces robust, neutralizing antibody responses that are stable for at least three months. medRxiv 2020; : 2020.07.14.20151126.

42 Pradenas E, Trinité B, Urrea V, et al. Stable neutralizing antibody levels 6 months after mild and severe COVID-19 episodes. Med N Y N 2021; 2: 313-320.e4.

43 City Executive Office, Urban Research and Statistics. Helsinki facts and figures 2019. https://www.hel.fi/hel2/tietokeskus/julkaisut/pdf/19_06_14_HKI-taskutilasto2019_eng_w.pdf.

44 Jones TC, Biele G, Mühlemann B, et al. Estimating infectiousness throughout SARS-CoV-2 infection course. Science 2021; published online May 25. DOI:10.1126/science.abi5273.

