## Supplementary figures for "High secondary attack rate and persistence of SARS-CoV-2 antibodies in household transmission study participants, Finland 2020"

**Supplementary Figure 1: Development and follow-up of IgG antibodies to SARS-CoV-2 nucleoprotein among confirmed cases, individual results**

Supplementary figure 1A: Asymptomatic cases

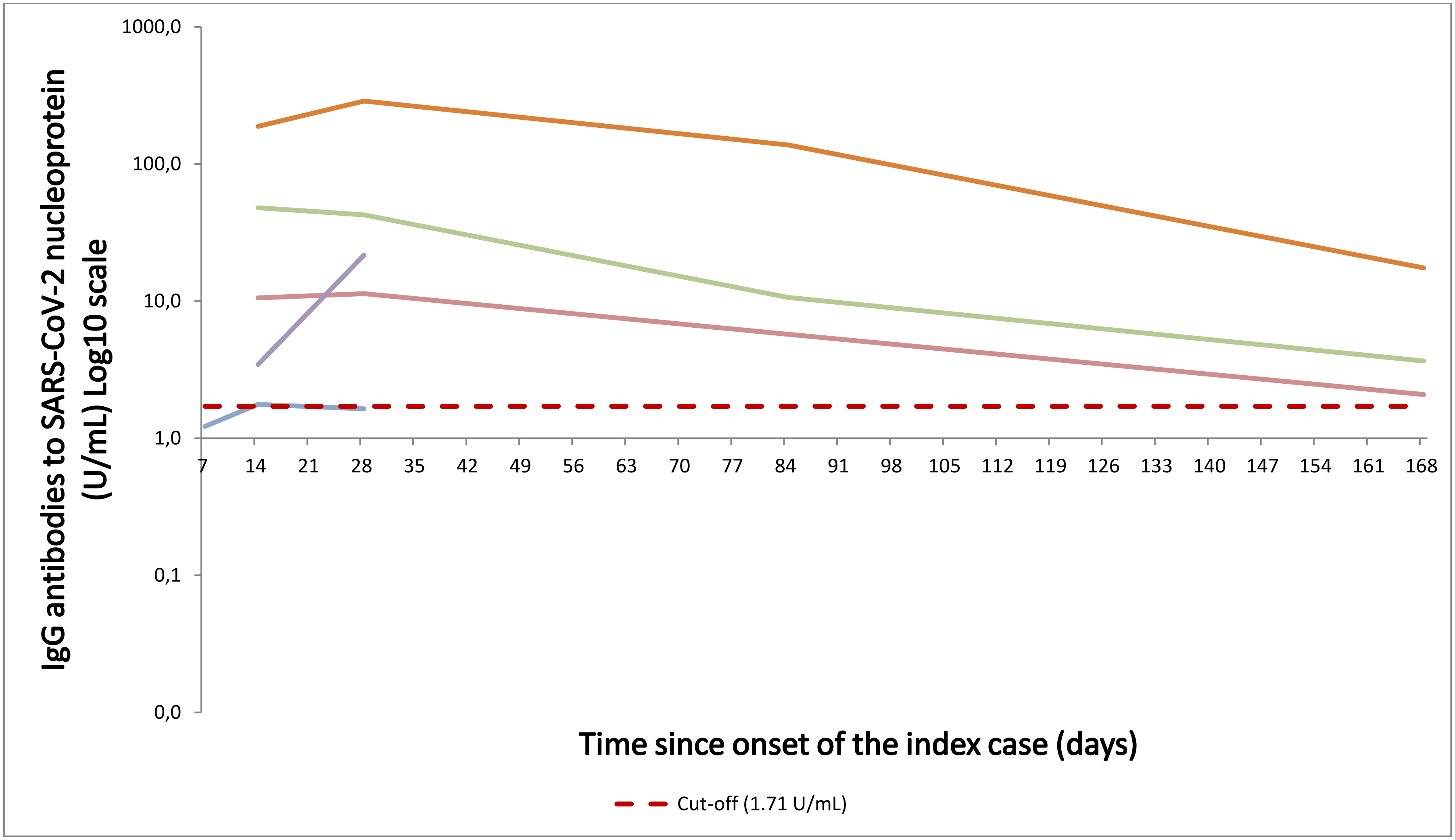

Supplementary Figure 1B: Cases with symptoms

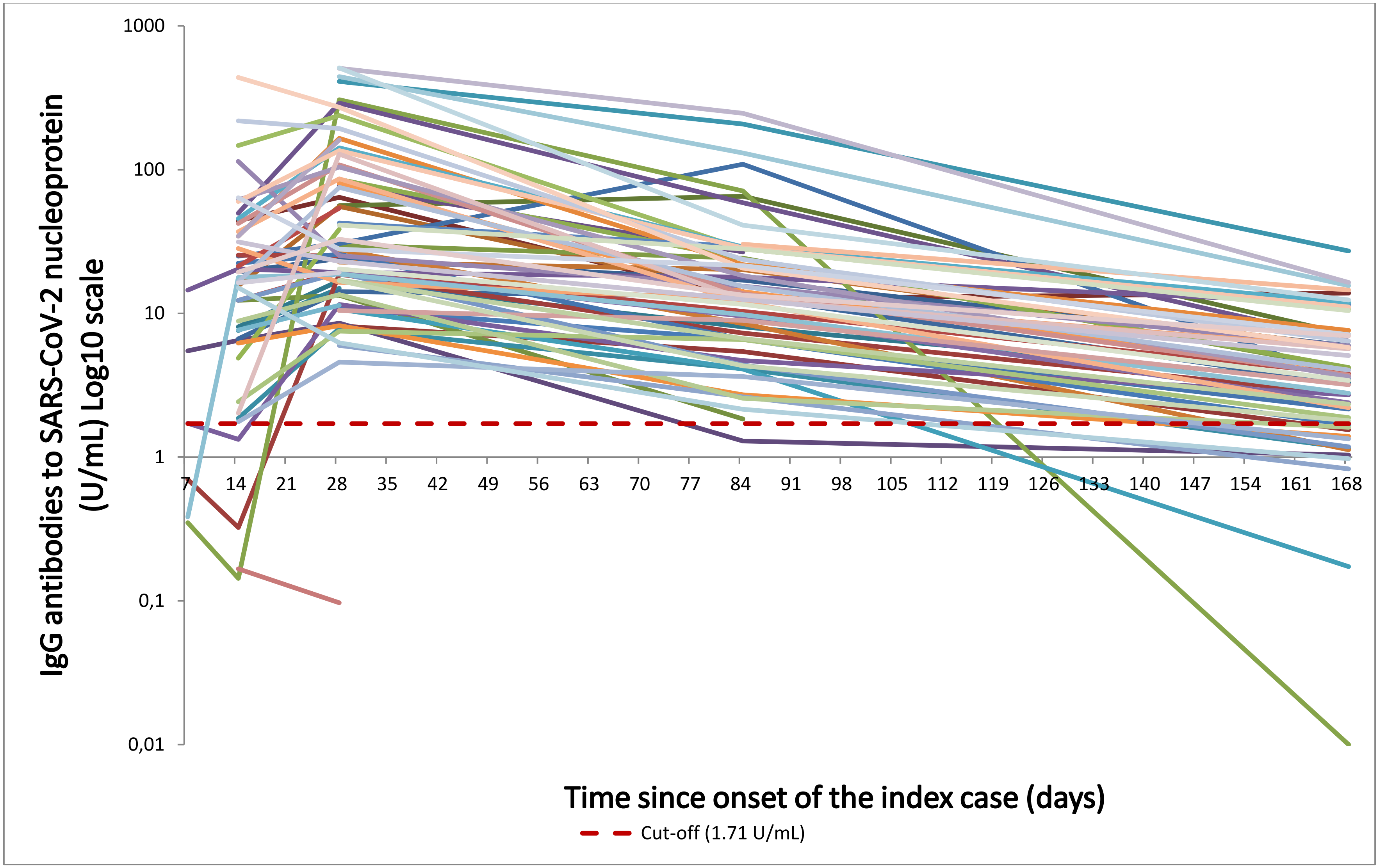

Supplementary Figure 1C: Cases requiring hospital care

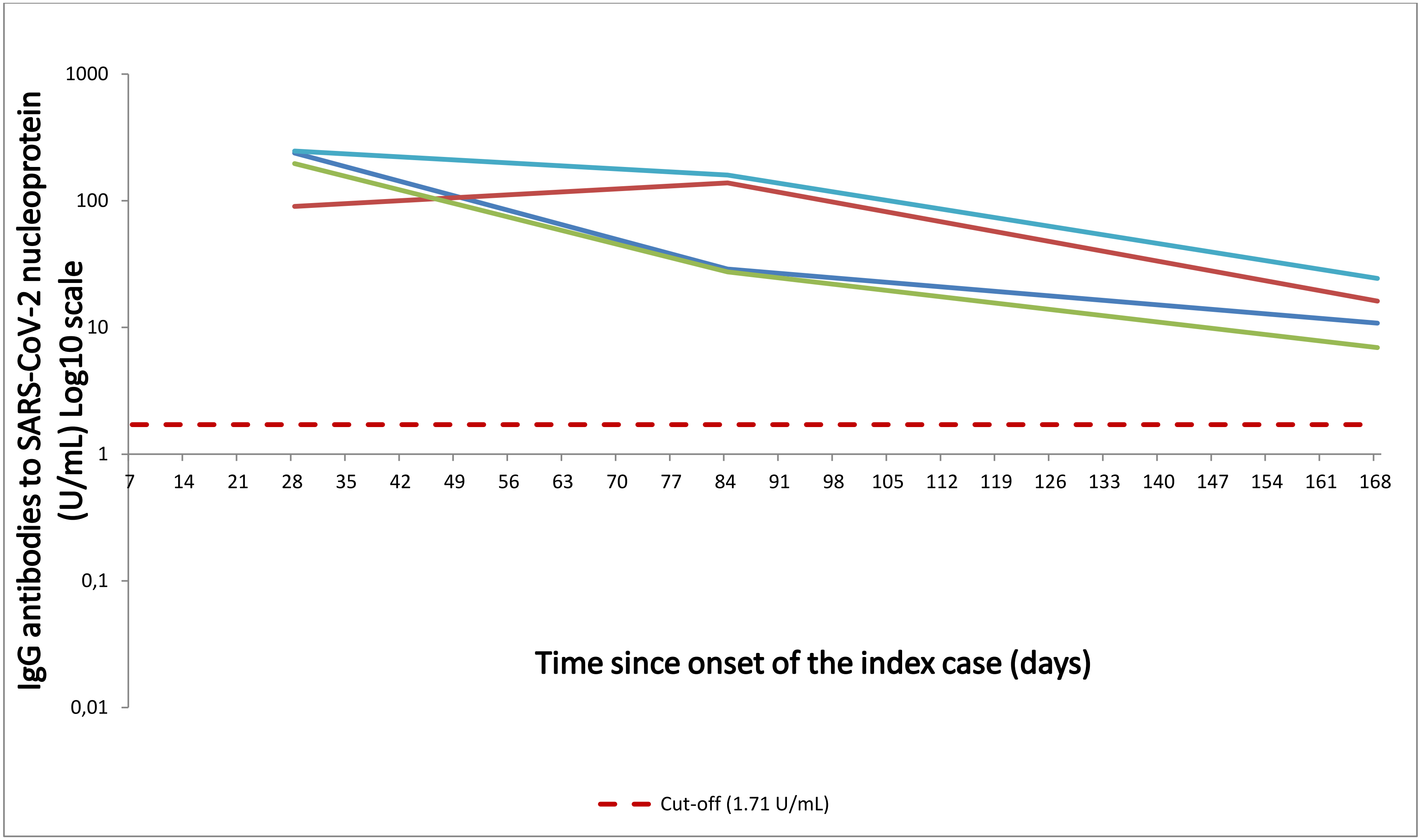

**Supplementary Figure 2: Trends in neutralization titers over time: Fin1-20, re-analysis of samples collected at 28 days, 3 months and 6 months in the same assay, geometric mean and 95% confidence interval.**

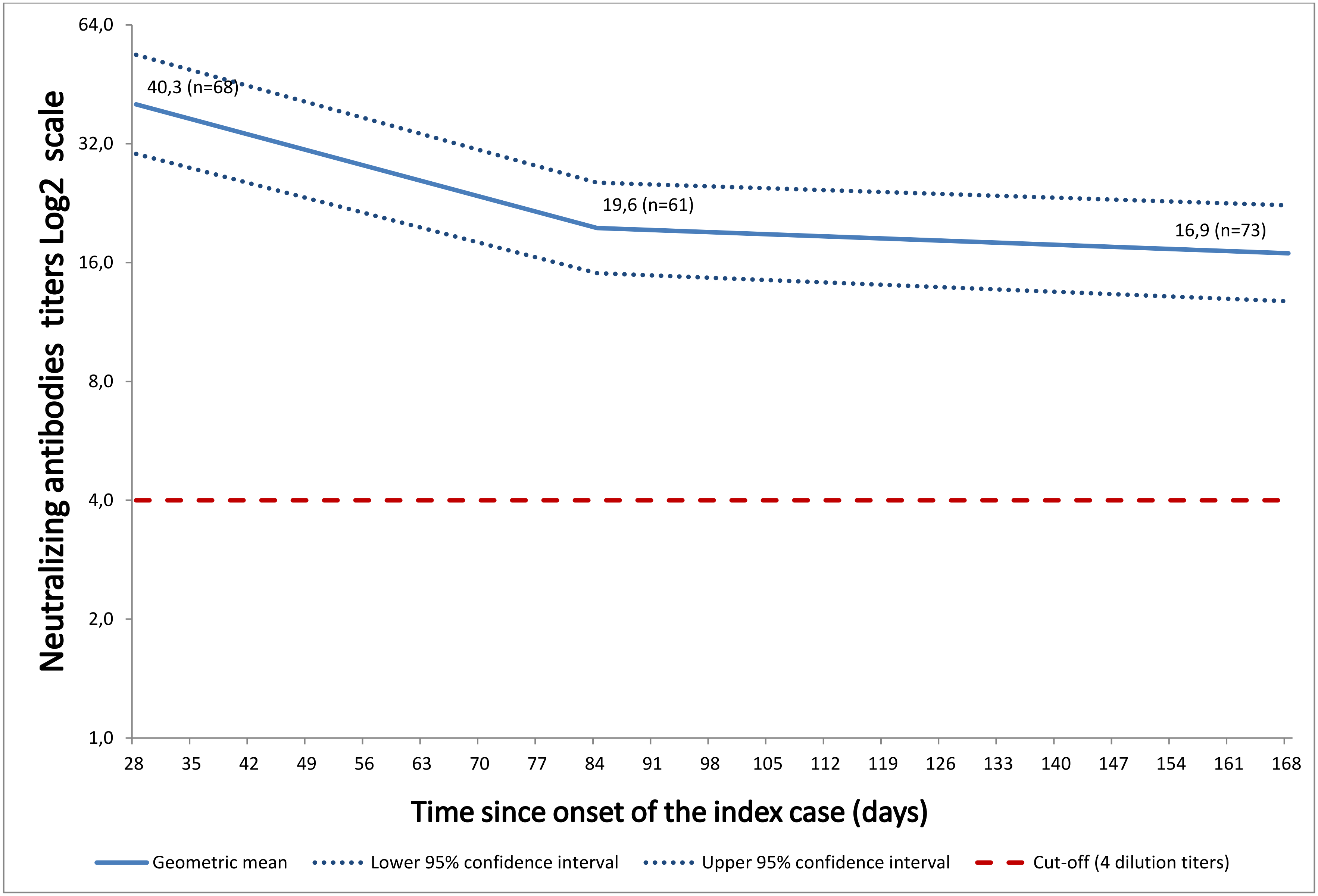

**Supplementary Figure 3: Trends in neutralization titers over time: Fin1-20, re-analysis of samples collected at 14 days, 28 days, 3 months and 6 months in the same assay, individual results**

Supplementary figure 3A: Asymptomatic cases

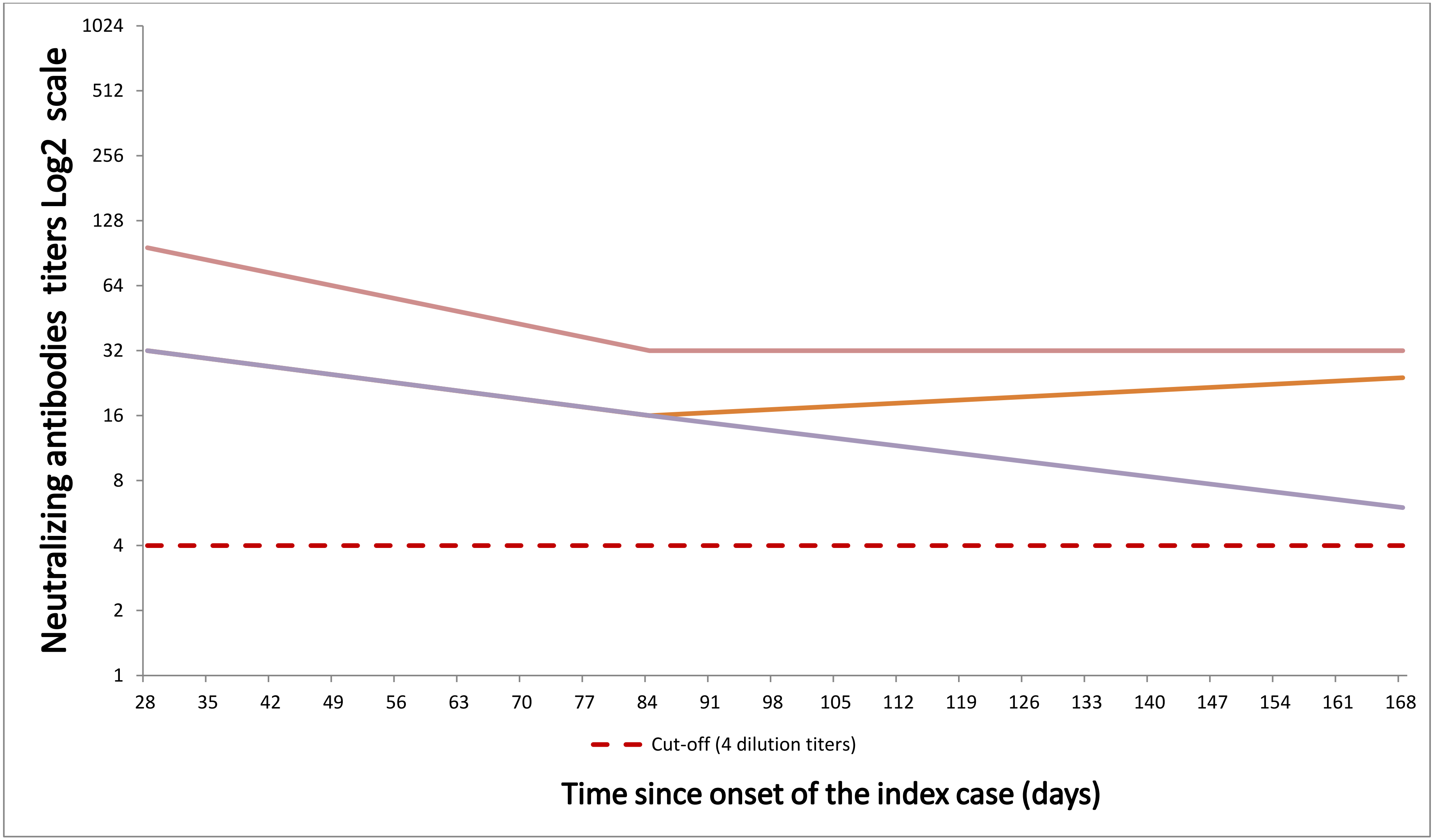

Supplementary figure 3B: Cases with symptoms

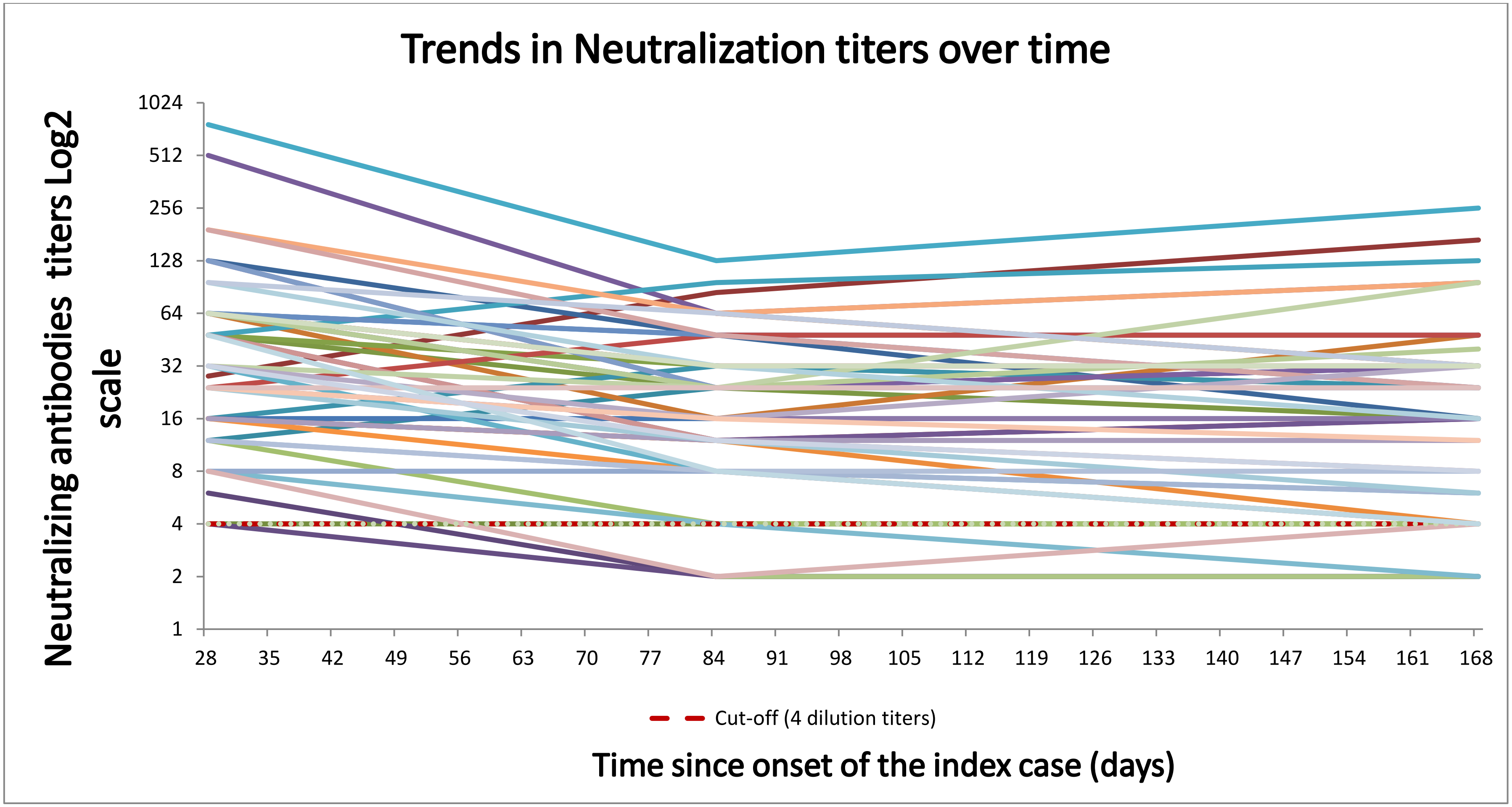

Supplementary figure 3C: Cases requiring hospital care

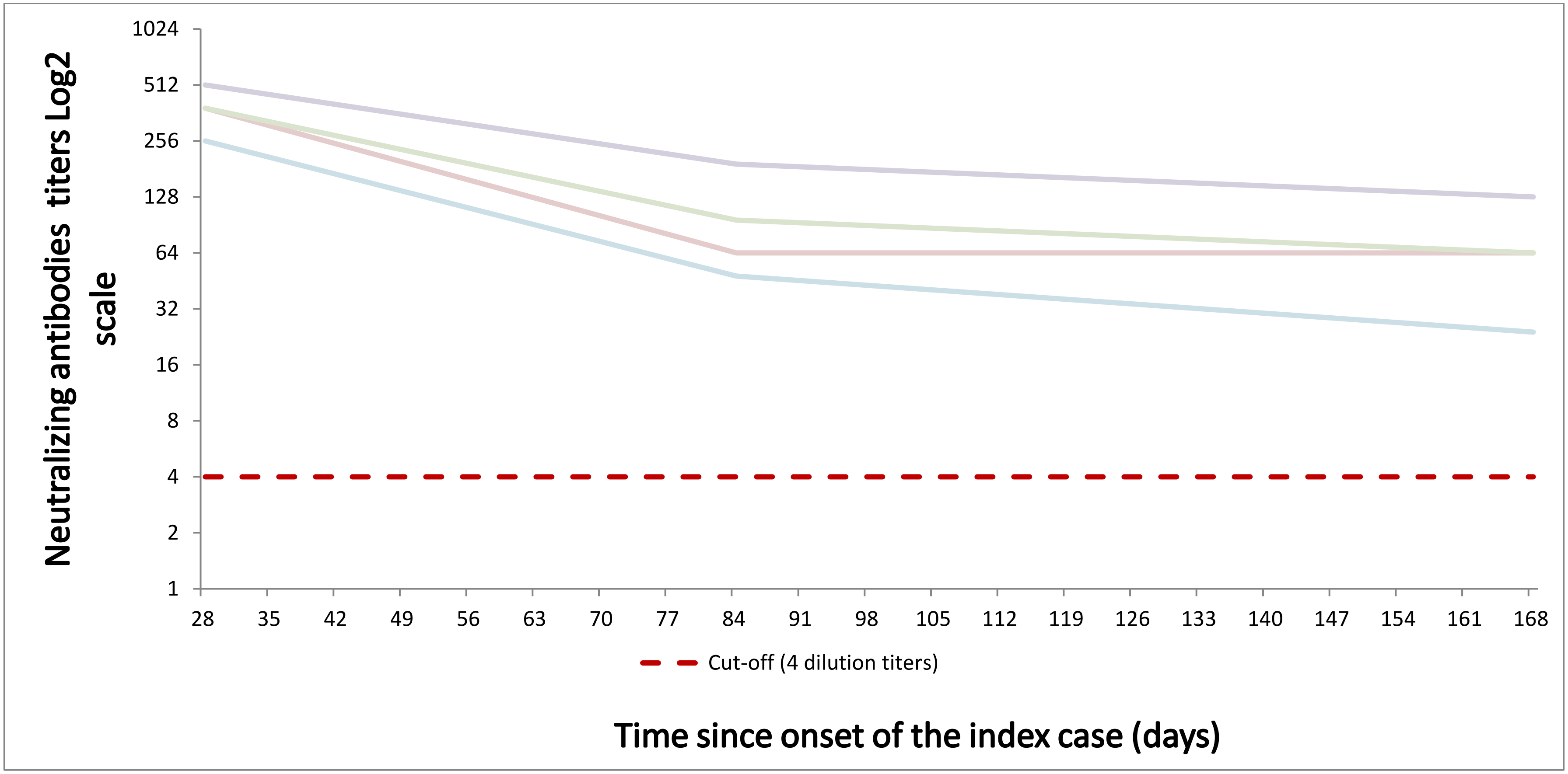
